# A novel epidemiological model for COVID-19

**DOI:** 10.1101/2020.07.23.20160580

**Authors:** Mauro Gaspari

## Abstract

COVID-19 is characterized by a large number of asymptomatic and mild cases that are difficult to detect; most of them remain unknown, still having an important role in the transmission of the disease, this makes the pandemic difficult to control. The purpose of this research is to develop an epidemiological model that allows to estimate the number of unknown/asymptomatic cases in a given area.

The SEIAMPR system, a novel simulation based model for COVID-19 is designed and implemented in Python. The intuition of the model is simple: about 80% of COVID-19 infected people evolve as asymptomatic or with a mild clinical course, many of them remain unknown to the authorities, some of them including those in critical conditions are eventually detected and classified as positive cases. The simulator reproduces this process using an adaptive method integrated with official data.

The simulator has been used for modelling the outbreak in 21 regions in Italy. The positive effects of lockdown policies are demonstrated: unknown active cases 12 days after the lockdown (March the 21th) ranged from 284101 to 374038, e.g. many more than all the official cases in Italy, reducing to 10213/20949 the reopening day. The number of unknown active cases at the beginning of June in the Lombardia region ranged from 6813 to 13390 demanding particular attention.

SEIAMPR is simple to tune and integrate with official data, it emerges as an up-and-coming tool for reporting the effect of lockdown measures, the impact of the disease on the population, and the remaining unknown active cases for evaluating the timing of exit strategies.

**Mathematics Subject Classification (2010)** MSC 68-XX

## 1 Introduction

The severity of COVID-19 and its high transmission rate required unpopular political decisions to control the spread of the virus in many countries, like weeks of lockdown and banning movement between the states or regions. The length of these unprecedented control strategies has caused immense loss of economic productivity, adverse social and psychological impacts on the population [1], [2].

In the situation which has arisen, it is essential for politicians to have available good justifications and concrete evidences for the adopted strategies, together with effective decision support tools for pursuing safe and ground-breaking exit strategies. Natural questions arise like: are the effects of policy measures observable? What would have happened without these measures? which is the real impact of the disease on population? How many were the unknown active cases before lockdown? and how many are the remaining unknown active cases?

It’s not surprising that due to the outbreak of the pandemic in the last months there has been increased research efforts on epidemiological models to provide answers to the above questions. Despite all these efforts, the epidemiological characteristics of pandemic COVID-19 are difficult to capture given the large number of asymptomatic and unknown cases, that are not reported in official data. As a result, the COVID-19 pandemic cannot be easily modeled with basic mathematical models, and more complex proposals like [3], [4] and [7] has proven necessary. These proposals include many variables and are difficult to tune and use, adapting them to different countries and regions. Moreover, they have been developed to answer specific questions, not all the possible ones [5].

In this paper, I present a novel epidemiological model for COVID-19 based on a simulated SEIR schema embedded within real data. Although, it is not designed as a tool for forecasting the development of the disease at the beginning of the outbreak, it emerges as an up-and-coming tool for reporting the effect of lockdown measures, the impact of the disease on the population, and the remaining unknown active cases for evaluating the timing of exit strategies. Most importantly, it is very simple to use and integrate with real data, providing a realistic estimate of the number of asymptomatic and unknown cases in a given area. To highlight its potential, I present a case study modelling all the Italian regions at the beginning of June 2020.

## 2 Modelling COVID-19 outbreak

Mathematical modelling based approaches [8] express relevant epidemiological knowledge through a set of variable and equations, which are solved and tuned to determine a function that represents the hypothetical evolution of a disease, plotting it on a computer. The models are usually classified with acronyms that represent the flow of patterns between the modelled compartments, for example SEIR stand for Susceptible, Exposed, Infected and Recovered. Mathematical models have been used as effective forecasting tools in many diseases extending the basic setting with other compartments [9]. For example, considering COVID-19 both basic models [6] and more complex models including asymptomatic [16] and age groups compartments [10] have been experimented in many countries.

### 2.1 The simulated SEIAMPR model

*SEIAMPR* (Susceptible Exposed Infected Asymptomatic Mild Positive Recovered) is not a mathematical but a simulation based model. The *Infected* compartment is organized in three categories of individuals, *Asymptomatic, Mild* unknown cases and official *Positive* cases in a given day. The *Recovered* compartment includes recovered cases from the three above categories. The *Exposed* compartment includes all the individual that have been exposed to the virus and will become infected after an incubation period.

The logical schema of the simulated outbreak is presented in Figure 1. Continue lines represent the flow of official data, dashed lines represent the flow of simulated data, the fine dashed line to the Deads compartment is not used in the model, is that of official data. The time series of current positive, new positive, infected, recovered and nasopharyngeal swab (NS) are needed to run the model, these are the only data that the model needs.

**Fig. 1.**
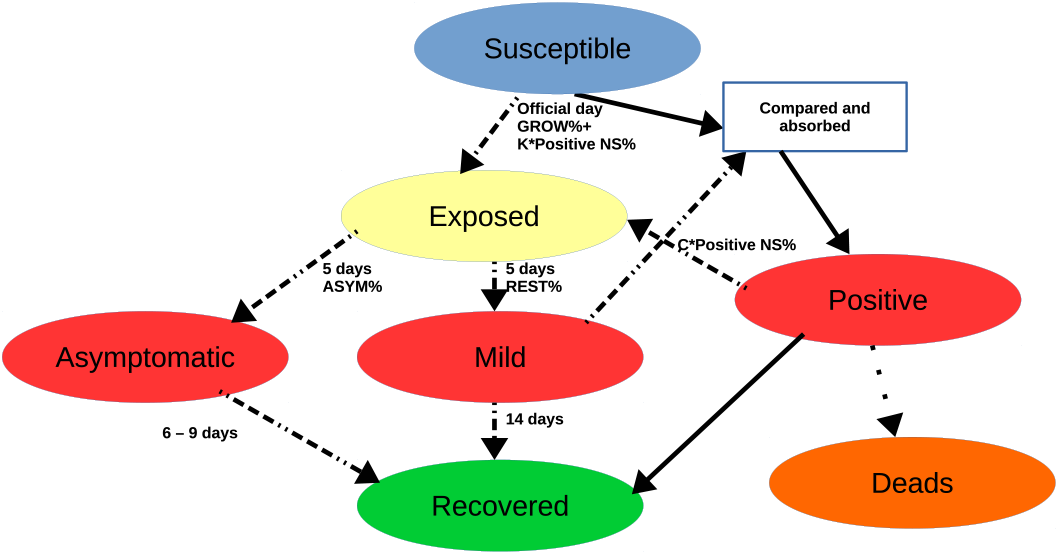
The SEIAMPR simulator schema: we use red for the three Infected categories, blue, yellow and green is used for susceptible, exposed and recovered respectively. Dashed arrows represent the flow of simulated data, continuous arrows represents real data. The main parameters of the simulator are associated to the arrows. All of them can be personalized to test different hypothesis.

The intuition of the model is simple: about 80% of COVID-19 infected people evolve as asymptomatic or with a mild clinical course, many of them remain unknown to the authorities, some of them including those in critical conditions are eventually detected with NS and classified as positive in official data. When an individual is exposed, after an incubation period, he/she starts with an asymptomatic or pre-symptpomatic, mild clinical course, successively some of the mild cases become more critical and they are eventually detected. This is the simulated epidemic process, the Positive compartment is those containing official data, it communicates the Mild compartment and the Exposed compartment. More precisely, new detected positives must belong from the Mild compartment that should contain a sufficient number of cases.

The simulator uses the official day grow percentage to determine the number of new exposed in the unknown Infected compartments each day. Apparently this choice is not accurate because the individuals that caused the grow percentage had been exposed to the virus several days before the day in which they were detected. However, the grow percentage is only one of the factors that determine the evolution of unknown cases in the epidemic model, what happens is that the curve is automatically adapted each day considering its relationship with official positives. More precisely, each day the new official positives are matched with the individuals which are at the end of the mild duration period, and the Mild compartment is adjusted to capture (removing them) all the new official cases. The set of Susceptible is modeled using an increasing factor and the NS day percentage, the higher is the NS percentage the more is the number of susceptible that become exposed in a given day.

Additionally, I assume a little percentage of exposed individuals that are infected by official positives after then they are detected. This percentage is multiplied by a factor that can be specified for each modelled area if needed. This factor is multiplied with the positive NS percentage of the day, to determine the new cases to load in the Exposed compartment.

The simulator assumes the following random variables and basic percentages that can be tuned for each modelled area, if more specific data are available.

– *ASYM* : 43.2% is the asymptomatic percentage, I used this proportion at the beginning of the simulation process, and at every stage, when new infected come up from the Exposed compartment. This percentage comes from the most important and thorough study on asymptomatic in Italy [11]; several studies around the world confirm a similar ratio [14].
– *NE* : 5 days is the average incubation period [15], [11]. I used a random generator based on wald distribution with mean 5.0 and shape 20, see Figure 2.

**Fig. 2.**
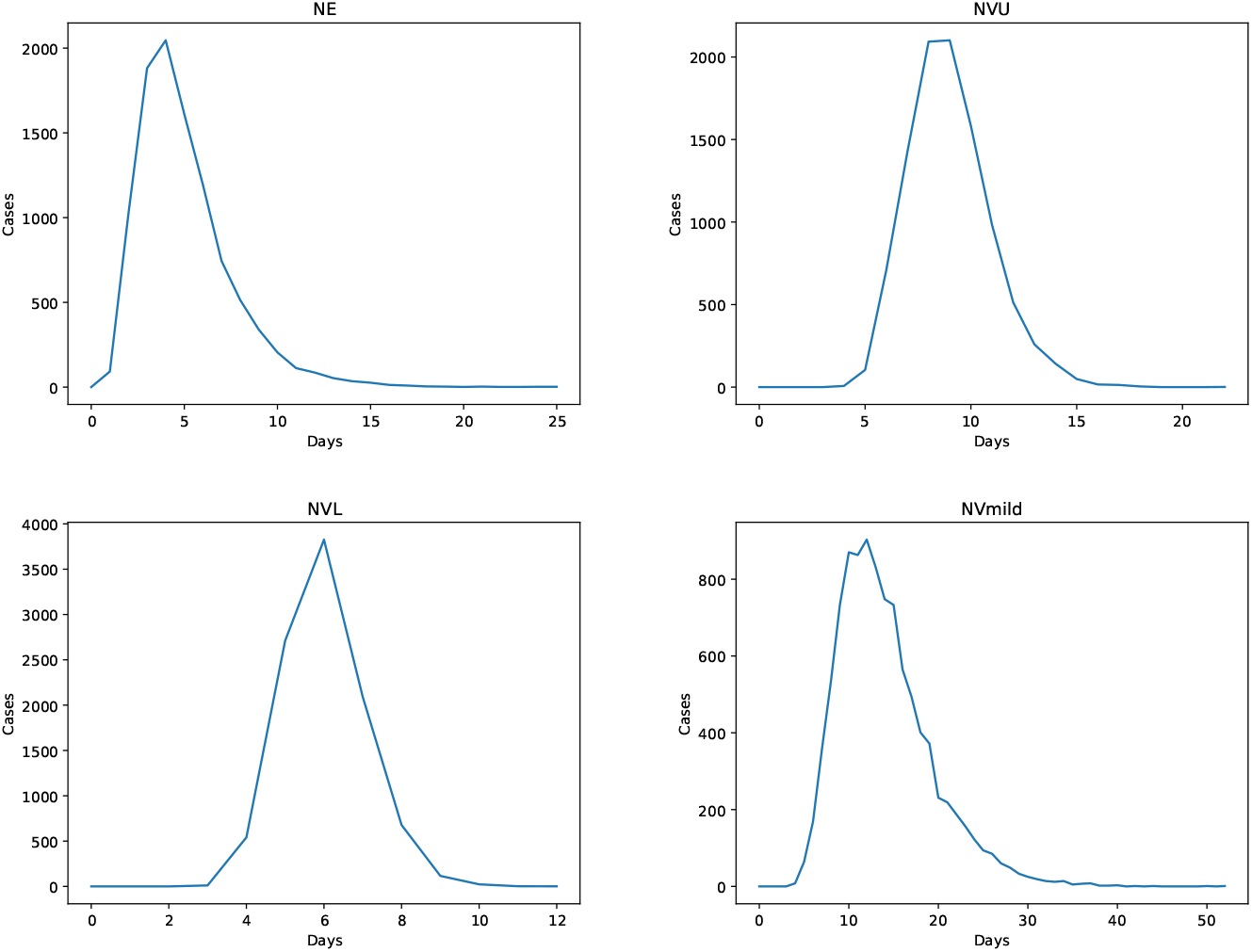
Distributions used for random generation of variables NE (5.0,20), NVU (9.0,200), NVL (6,0,200), NVmild (14.0,100). The diagrams are plotted considering 10000 cases. All of them are based on the binomial-inverse (wald) distributions using the above parameters for mean and shape.
– *NVU* : 9 days for the asymptomatic duration period [13], [11]. I assumed this as an upper bound for asymptomatic course. I used a random generator based on wald distribution with mean 9.0 and shape 200, which generates values ranging from 3 to 21 days, see Figure 2.
– *NVL*: 6 days is the lower bound for the asymptomatic duration period [13]. It is not misleading to assume this shorter interval: although, the average value reported in [11] is higher, it is likely that some asymptomatic cases where not detected, for example children, because they cleared the infection between the two surveys. I used a random generator based on wald distribution with mean 6.0 and shape 200 as above, which generates values from 3 to 12 days, see Figure 2.
– *NVmild* : 14 days is the average duration period I assumed for symptomatic mild cases [17]. I used a random generator based on wald distribution with mean 14.0 and shape 100, which generates values from 4 to about 50 days, see Figure 2. This distribution also models the time period in which these cases are detected and migrate to the official Positive compartment (details are presented in the next section). More precisely, some of them will be detected by NS and moved to the Positive compartment, other will remain unknown and discharged at the end of the random generated period.

### 2.2 The simulation process

The SEIAMPR simulation process is implemented in Python and it is organized as follows: To start a simulation the set of initial cases must be estimated, these cases are considered infected (see Section 2.3). Infected cases are loaded in the Asymptomatic and Mild compartments and the discharge dates using the above distributions are set for each case in the compartments.

For each day the simulator executes the following steps:

1. It computes the day grow percentage *GROWP* which it is used to find the number of individuals that have been infected by unknown cases (in the Asymptomatic and Mild compartments). Let *CP* and *dayP* be respectively the current official positive cases and the new positive cases of the day, and let *NSdayP* be the positive NS percentage of the day, *GROWP* is obtained as follows:

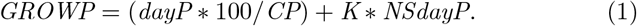

where *K* is an increment factor that can be a constant, for example 0.2, or a variable coefficient defined by a function. Intuitively, I hypothesize that unknown cases grow like known ones considering an addend that depends on the positive NS percentage. The higher is this percentage, the more is the number of unknown infected cases. The positive NS percentage of a day *NSdayP* is estimated removing from the total number of NS the number of recovered cases in the same day, assuming at least one positive NS for each of them.
2. It computes the number of individuals *NPOS* that have been infected by official positive cases. They are determined using the following formula:

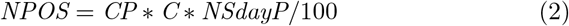

where *C* is an increment factor that can be a constant, for example 0.1, or a variable coefficient defined by a function. The result depends from the positive NS percentage too.
3. Infected cases individuated in the previous two steps are first loaded in the Exposed compartment, indeed, they will become infected only after an incubation period. The simulator sets the discharge dates for them using *NE* as mean and the above distribution (Figure 2).
4. If in the same day there are cases that are at the end of the incubation period, they are moved in the Asymptomatic or Mild compartments using the *ASYM* percentage to break them up into the two sets.
5. Then the simulator updates the Asymptomatic and Mild compartments removing the cases that recover according to the registers, and it removes the new infected (i.e. *dayP*) from the Mild compartment, which are moved to the official Positive compartment.
6. When necessary, if the number of new unknown cases of the day is not enough, e.g, it is less then *dayP*, the simulator removes from the Mild compartment previous unused cases (as a default 7 days before) and exceeding cases are deleted from the recover register of mild cases in the next days (as a default these exceeding cases should be absorbed 4 days after). Intuitively these are the cases that have been discovered by NS, thus they are removed from the simulated sets and become part of official data. With this extended interval Mild cases can be discovered about 2 to 20 days after infection. Using this method the unknown cases curve is automatically adapted to the evolution of official positive cases.

All the parameters of random variables, the values of increment factors and intervals used in the simulator can be tailored when new evidences arise or to test how the outbreak evolves in particular situations and hypothesis. For example, considering a different percentage for asymptomatic during the summer, a different duration period for mild cases or a longer incubation period.

The described simulation process is extended with a bootstrap mechanism to model the first days of the epidemic, and with an accumulator based mechanism for the last days.

The simulator takes as input an estimated number of infected individuals that are loaded in the Mild compartment all together. As a consequence of this initialization procedure in the first days of the simulation the mild register is almost empty, because the assumed distribution generates only a few cases at the beginning. Thus, the first days there are not enough mild cases loaded in the register to justify the new positives; to fill them the initial set of infected must be overestimated. To solve this bootstrap problem, I introduced an initial buffer of infected cases that is used at the very beginning of the process when the Mild register is almost empty. These cases are directly moved to the official Positive compartment.

On the contrary, at the end of the epidemic curve the number of unknown cases becomes smaller and smaller, and the *GROWP* percentage may generate 0 new unknown cases. When this happens unused cases are stored in an accumulator which is emptied only when new cases are generated. The same solution is also used for individual infected by official positives.

### 2.3 Exploiting the simulator with real data

Given the positive, new positive, infected, recovered and NS time series, the simulator needs as input the number of infected individuals at the beginning of the outbreak, which is the day in which the first infected person was detected and reported in official data. These individuals are many more than the official cases, see for example [18] or [12]. However, it was difficult to figure out a uniform estimation method working well for all the regions of Italy, given that the situation of them was not uniform at the beginning of the outbreak.

Considering that this model was developed at the end of May, when all the Italian regions had a downward trend, a semi-automatic tuning procedure turned out to be effective. It is based on the insight that the simulator needs a sufficient number of unknown cases to justify and absorb the new positives reported in official data in each day.

The procedure starts hypothesizing an initial (small) number of unknown cases in the first day of the outbreak in a given area. The simulator is activated with this number as an input to check if it is able to generate all the official positive cases until a given day, taking them from the Mild compartment. This is obtained using the auto-adaptive method introduced above: if there are N official new positives in a given day the simulation process should be able to generate at least N cases in the Mild compartment in the same day. If this condition holds these cases are moved to the official Positive compartment and removed from the Mild one. Otherwise, if the cases generated in that day are not enough, the simulator considers cases generated some days before and some days after. The interval is specified by two variables *BAPPR* (default 7) and *FAPPR* (default 4). If this interval is enlarged the number of initial cases decreases, but the precision of the tuning procedure gets worse.

If the simulator is not able to generate all the official cases in the assumed interval of days an overflow message is printed, this means that the number of initial cases was not sufficient, and that it must be increased. This process is iterated until the simulator is able to generate all the official cases without overflow messages.

The intuition beyond this method is that the number of infected individuals at the beginning of the outbreak should be enough to support all the current positive cases, passing the epidemic peak.

## 3 Results

I tested the SEIAMPR simulator using the data provided by the Italian Protezione Civile Deparment [19] analysing the outbreak in the 21 regions of Italy. The simulator was tuned the 2nd of June (the day before the movements between regions were reinstated) using the method presented in the previous Section (2.1). I used a 5 days duration period for asymptomatic in the tuning procedure and the default setting for parameters and variables described above. This can be taken as a quite optimistic lower bound for all the modelled regions.

Considering the *C* and *K* increment factors, I used two linear functions ranging respectively from 0.2 to 0.1 for *C* and from 0.12 to 0.25 for *K*. Intuitively, *C* decreases over time because the ability to isolate known positive has improved, while *K* increases because the number of susceptible has decreased. Thus, the weight of the same NS percentage is higher in the last days of the curve, with respect to the first days. This setting gave reasonable results for all the regions of Italy.

The simulator is executed 11 times for each region and the average values are reported for each region. The generated epidemiological curves for unknown cases considering an average 6 days duration period for asymptomatic and an average 9 days duration period are presented in Figures 3, 4 and 5, for north, center and south of Italy respectively. The figures show that unknown cases are many more of the official ones.

**Fig. 3.**
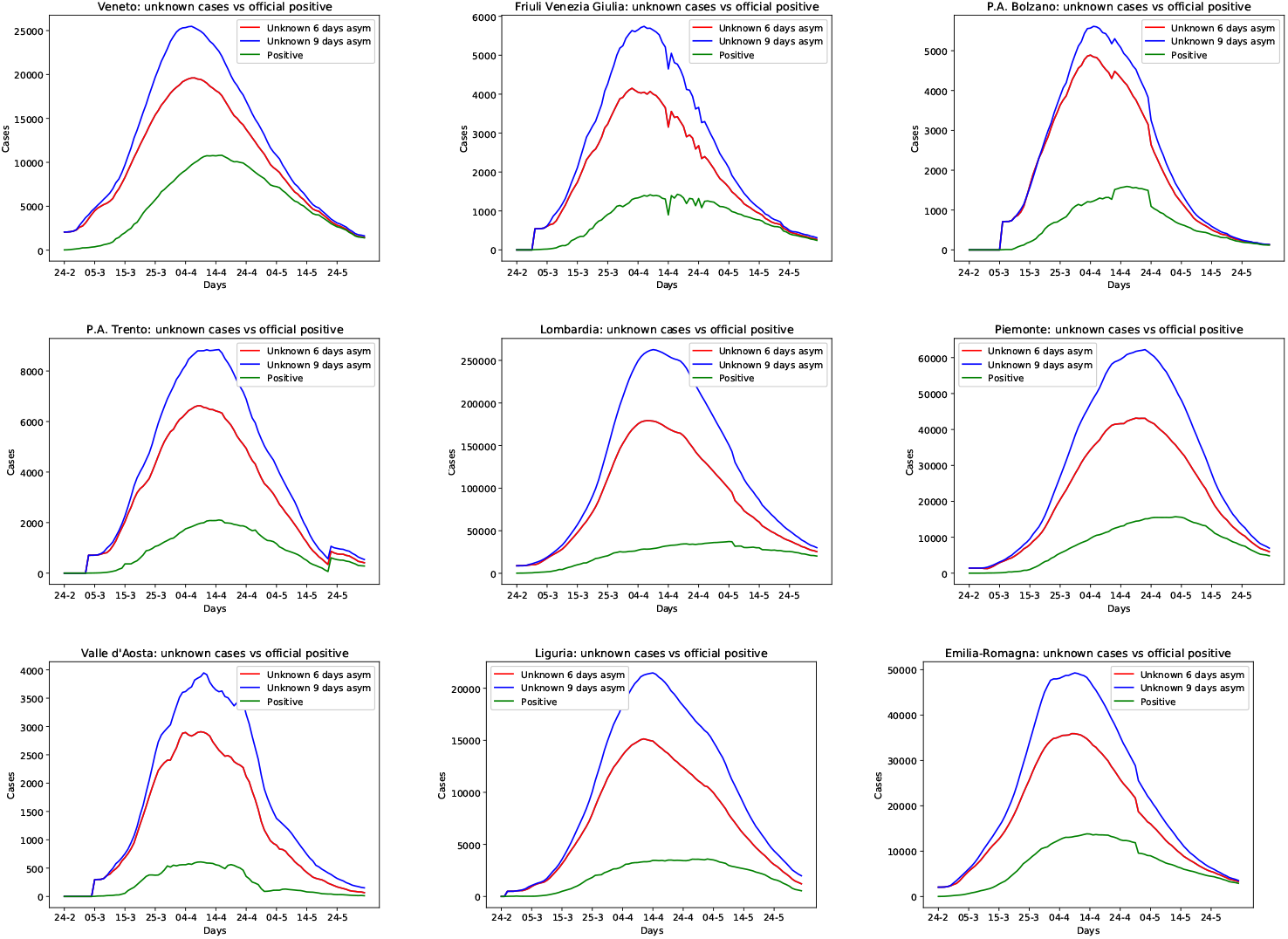
Epidemiological curves for unknown cases of the regions in the north of Italy. Green is used for official cases, while red and blues are used for unknown cases assuming 6 or 9 days asymptomatic duration period.

**Fig. 4.**
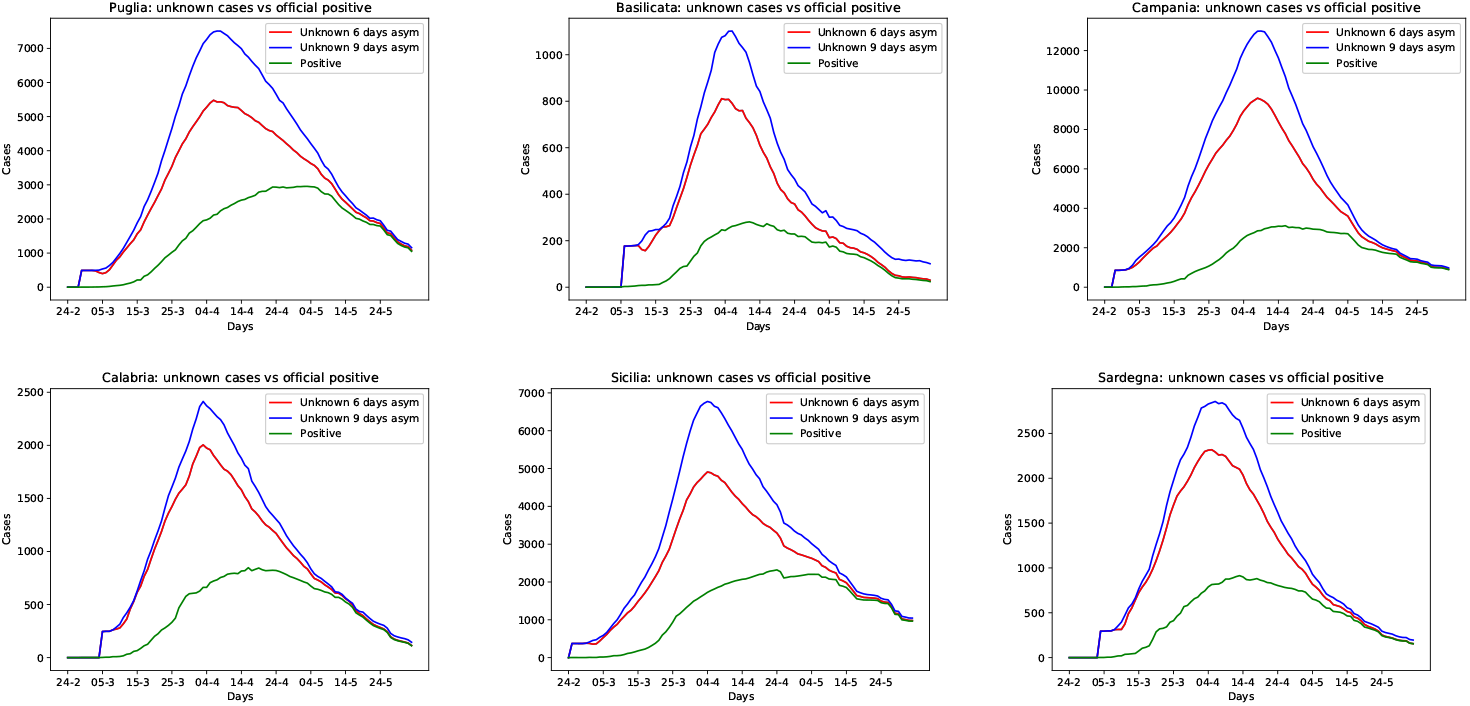
Epidemiological curves for unknown cases of the regions in the south of Italy. Green is used for official cases, while red and blues are used for unknown cases assuming 6 or 9 days asymptomatic duration period.

**Fig. 5.**
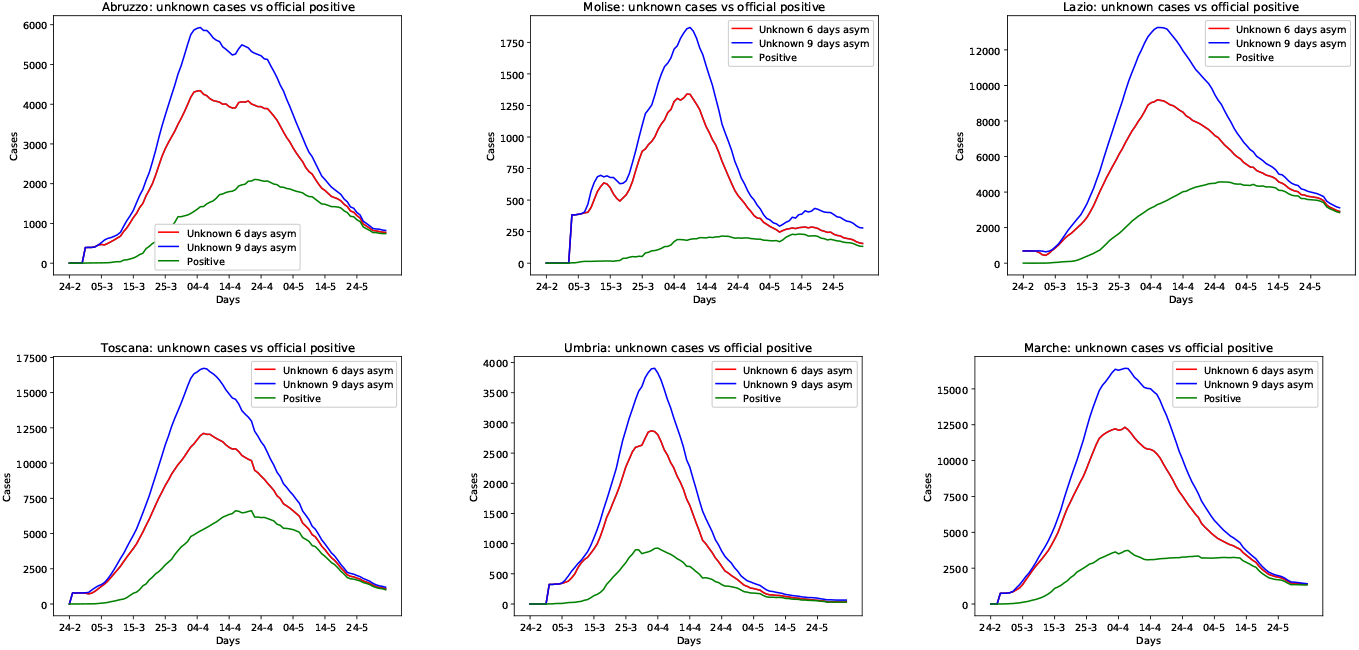
Epidemiological curves for unknown cases of the regions in the center of Italy.. Green is used for official cases, while red and blues are used for unknown cases assuming 6 or 9 days asymptomatic duration period.

Observing how the simulator works, I can do several considerations. In some of the regions the gap between unknown and official cases is larger with respect to others, in general this indicates that the adopted policies for controlling the disease were less effective. For example, this is the case of region Lombardia. However, there is a factor that depends on the parameters which are the same for all the Italian regions, so in some cases this statement may be misleading. Several factors should be considered together for a finer evaluation specific to each region. Nevertheless, the model provides an approximation of the pandemic that can be useful for many considerations.

The main results of the experiment are summarized in Table 1. This table presents the total number of cases and the unknown cases for each region compared with official ones. It also presents the number of unknown active cases the 21st of March (12 days after the lockdown that was the 9th of March) and the number of unknown active cases the 2nd of June 2020, the day before reopening movements between regions. On the one hand, the positive effect of the lockdown policy is clearly reported, indeed the estimated unknown active cases after the lockdown ranged from 284101 to 374038, e.g. more than all the official cases of Italy until the 1st of July. This number also includes the exposed individuals still in incubation period, that the simulator allows to report. The ranges of the best and worst cases for Italy considering the 11 simulations are respectively: 284492/373483 and 285296/380311 indicating that the simulation process converge. On the other hand, the number of unknown active cases at the beginning of June in the Lombardia region: 6813/13390 demanded particular attention, especially considering the possible effects on the other regions where the epidemic was almost at the end and in the tourist areas.

**Table 1.** Summary for Italy the 2nd of June 2020: the Table shows the total number of cases, the official cases, the total number of unknown cases, the number of unknown active cases 12 days after lockdown and the remaining unknown active cases, providing the estimates for both 6/9 asymptomatic duration periods.

| Region | Total Cases |  | Unknown | Active Cases |  |
| --- | --- | --- | --- | --- | --- |
|  | Total | Official |  | Lockdown | 06-02-20 |
| Abruzzo | 11867/16067 | 3249 | 8618/12818 | 3318/4320 | 44/89 |
| Basilicata | 1650/1998 | 399 | 1251/1599 | 602/699 | 4/58 |
| Bolzano | 12974/14318 | 2598 | 10376/11720 | 4445/4712 | 13/14 |
| Calabria | 4887/5746 | 1158 | 3729/4588 | 1981/2321 | 4/17 |
| Campania | 25445/33358 | 4809 | 20636/28549 | 8367/10700 | 23/94 |
| Emilia-Rom. | 102737/137680 | 27828 | 74909/109852 | 28710/37443 | 409/741 |
| Friuli VG | 11591/15127 | 3276 | 8315/11851 | 3853/4888 | 20/71 |
| Lazio | 22271/33097 | 7743 | 14528/25354 | 7145/10212 | 78/240 |
| Liguria | 52150/75321 | 9734 | 42416/65587 | 9620/12399 | 965/2080 |
| Lombardia | 648633/938735 | 89205 | 559428/849530 | 135254/181675 | 6813/13390 |
| Marche | 35238/45204 | 6734 | 28504/38470 | 10615/13497 | 65/90 |
| Molise | 4000/4846 | 436 | 3564/4410 | 1289/1499 | 20/115 |
| Piemonte | 133766/196262 | 30715 | 103051/165547 | 23101/30658 | 1359/2826 |
| Puglia | 12697/17496 | 4498 | 8199/12998 | 3988/5318 | 34/109 |
| Sardegna | 5536/6767 | 1357 | 4179/5410 | 2051/2504 | 4/19 |
| Sicilia | 10732/14550 | 3447 | 7285/11103 | 3559/4727 | 18/58 |
| Toscana | 29219/39916 | 10117 | 19102/29799 | 9861/13322 | 50/189 |
| Trento | 20077/26621 | 4432 | 15645/22189 | 5131/6372 | 138/286 |
| Umbria | 6307/7717 | 1431 | 4876/6286 | 2699/3306 | 5/34 |
| V. d'Aosta | 9676/12591 | 1187 | 8489/11404 | 2496/2953 | 94/221 |
| Veneto | 44925/57743 | 19162 | 25763/38581 | 16016/20513 | 53/208 |
| Italy | 1206378/1701160 | 233515 | 972863/1467645 | 284101/374038 | 10213/20949 |

The total number of cases estimated by the simulator in Italy: 1206378/1701160 is 5 to 8 times larger than the cases reported in official data (the worst case is 1210397/1728823). Given that the simulator generates the minimum number of unknown infected that is sufficient to support the official cases curve, this number can be considered an indicative lower bound for the real number of infected in Italy. This result is confirmed by the results of recent serological tests. For example in Lombardia total cases ranges from 648633 to 938735, e.g. more than a factor of 10 with respect to the official ones.

### 3.1 The Role of Asymptomatic

Observing Figures 3, 4 and 5, I can do some considerations about the role of asymptomatic in the epidemic. If the average asymptomatic duration period increases the number of total cases increases too, intuitively because the probability that they infect more people is higher. Also, if the length of the average asymptomatic duration period is lower, the length of the epidemic curve decreases. This means that policies that aim to boost the immune response of the population, like outdoor activities, could have a positive impact in controlling the outbreak. A shorter average duration period for asymptomatic could be an effect that occurs during the summer.

If the percentage of asymptomatic is higher with respect to the 43.2% assumed for this studio, the number of unknown cases needed to generate the official positives grows in a consistent way. This effect is illustrated by an experiment considering Veneto and Calabria regions presented in Figure 6, where the basic setting is compared with asymptomatic percentages of 60% or 70%. In Calabria unknown cases ranged from 3729 to 4703 (60%) or 6467 (70%), and in Veneto ranged from 25736 to 40138 (60%) or 60886 (70%) considering a 6 days average duration period. Therefore, if the percentage of asymptomatic effectively increases during the summer, behind an apparently small number of official cases, there could be hidden a consistent number of asymptomatic individuals. If this hypothesis is truth, a rapid uncontrollable rise of the outbreak would be possible in autumn, due to a considerable number of cases that gradually become more critical.

**Fig. 6.**
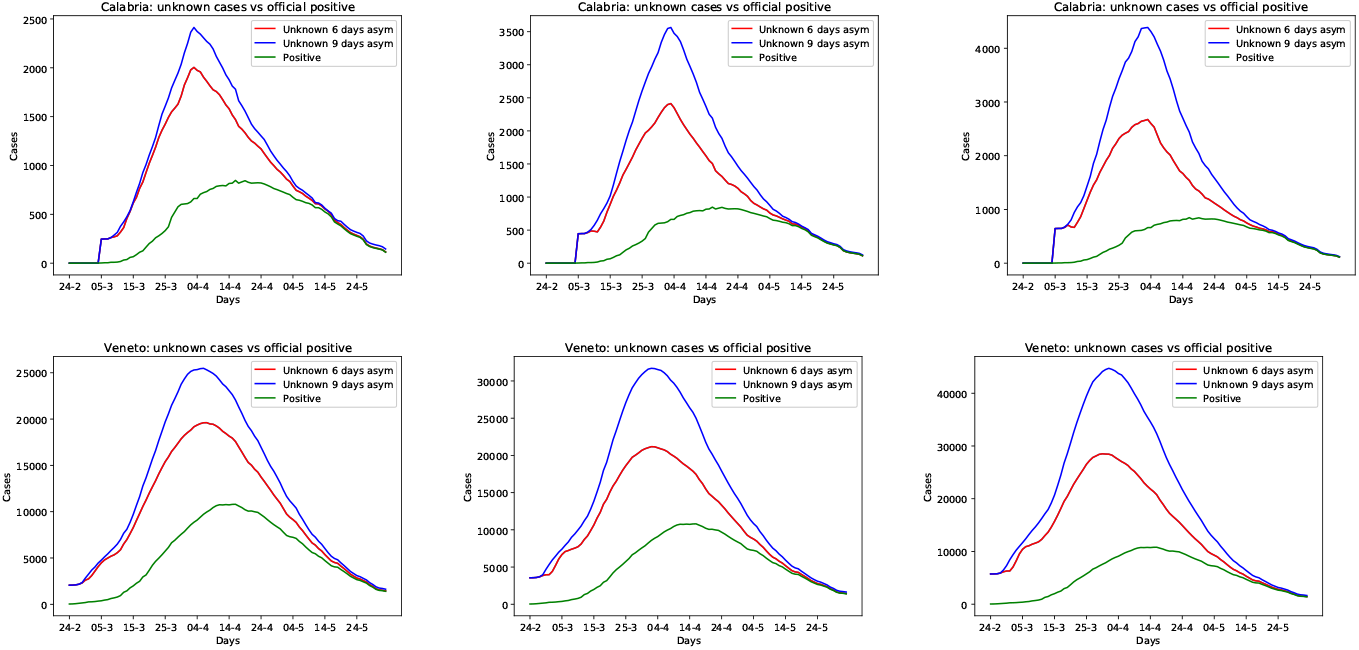
Calabria and Veneto regions: changing the asymptomatic percentage from 43.2% to 60% and 70% from left to right.

Another consideration concerns the ability to detect and isolate asymptomatic with adequate trace and screening strategies. In Figure 7 the impact of reducing the asymptomatic percentage is illustrated for Campania, Emilia Romagna and Lombardia regions, starting from the basic setting of 43.2% (on the top) to 20% (below). Considering Campania this change drastically reduces the total number of cases from 25445 to 9760, possibly this indicates that the results obtained with the basic setting for Campania is overestimated, if many asymptomatic were detected and isolated. Also in Emilia Romagna and Lombardia the number of cases is reduced, but the proportion is not so high.

**Fig. 7.**
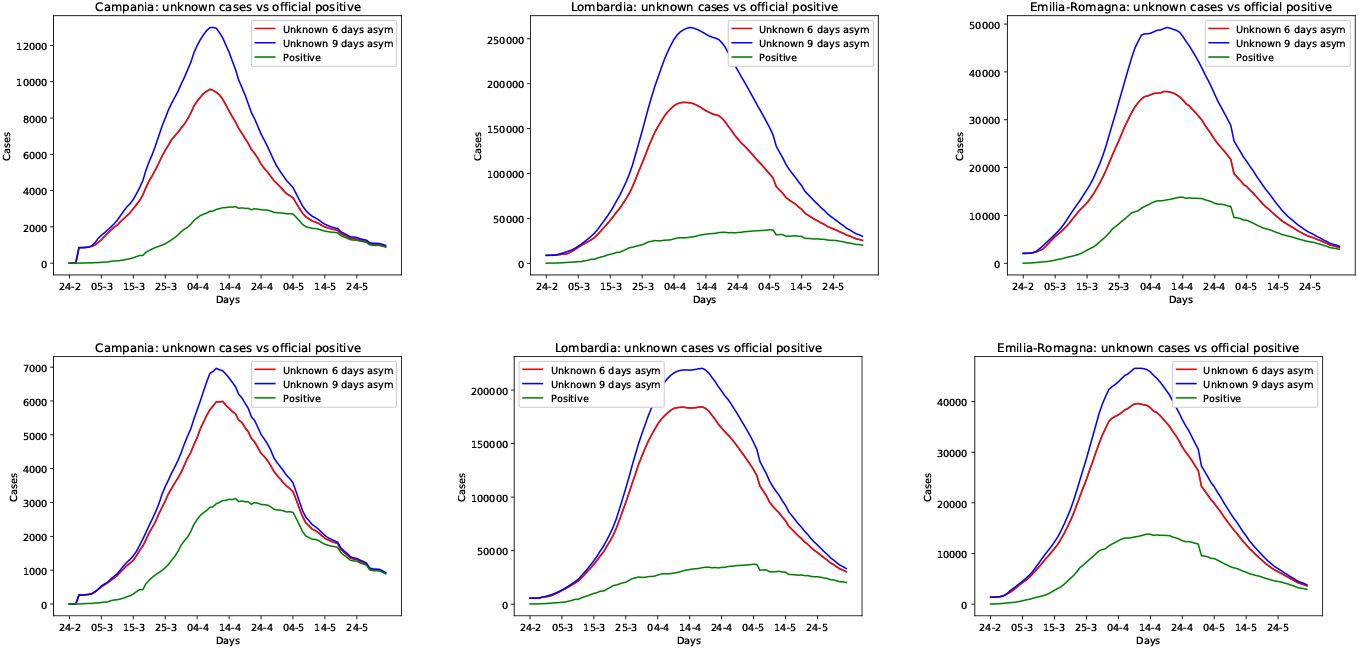
Reducing the percentage of unknown asymptomatic from 43.2% (on the top) to 20% (below) reduces the total number of cases: the Campania, Emilia Romagna and Lombardia regions.

### 3.2 Using death-based estimate for upward tuning

The simulator can be also initialized using estimations of infected people obtained with different methods. For example the automatic, semi-parametric estimation method developed by L. Fenga for all the regions of Italy [12].

Here, I will show how this method can be effectively used for tuning the SEIAMPR system. The benefit of this additional methodology are twofold: First, it provides an upward tuning tool that can be used at the beginning of the epidemic, while the semi-automatic tuning procedure illustrated in Section 2.3 only works in a downward setting, when the epidemiological curve has passed the peak. Second, if the result are compatible, it can be considered as a further assessment for the presented simulation based method.

I considered the four regions having the larger number of cases reported in official data which are: Lombardia, Emilia-Romagna, Veneto and Marche, comparing the total number of cases computed by SEIAMPR with the intervals reported in [12] for the infected at March the 12th, the results are shown in Table 2.

**Table 2.**
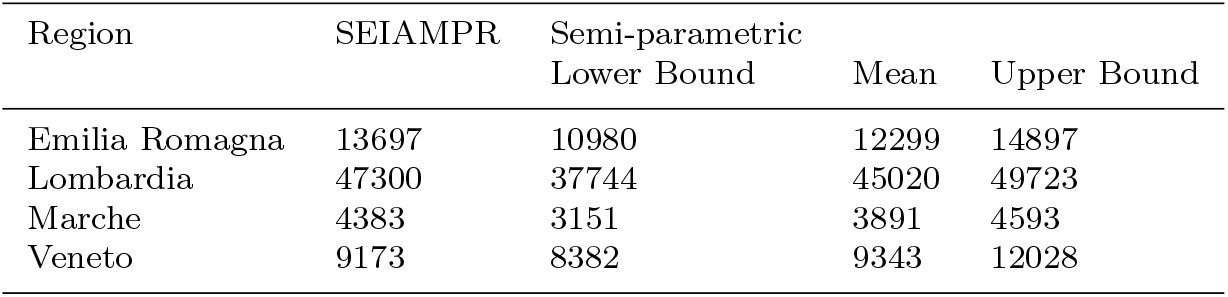
Total cases in Italy the 12th of March 2020: SEIAMPR vs Fenga’s automatic Semiparametric estimation method considering the Lower Bound, the Mean, and the Upper Bound.

The number of cases that the simulator computes for these regions is in the ranges of the automatic semi-parametric method intervals, while the simulator generates more cases for the other regions. This effect possibly depends from the low number of cases in the other regions the 12th of March. Indeed with a larger number of data statistical methods provide more accurate results.

Tuning the SEIAMPR simulator using the upper bound of the automatic semi-parametric method gave good results, compatible with those obtained the 2nd of June by the semi-automatic method presented in Section 2.3. This means that the SEIAMPR model can be effectively used at the beginning of an epidemic, providing an estimation of unknown cases also in a growing trend.

### 3.3 The last weeks

As the last point of this studio, I present the evolution of the Lombardia region, that was the most critical one at the beginning of June, simulating the epidemic until the 1st of July 2020. The setting used the 2nd of June was not enough to support all the official cases. However, updating the initial set of infected from 9100 to 9500, was enough to run the simulator successfully. With this setting the simulator computes for the 12th of March a number of infected compatible with the upper bound of the automatic semi-parametric method (see Table 2).

The results are reported in Figure 8 which presents an updated estimate for the 2nd of June and the last estimate at the beginning of July. Assuming an higher number of initial cases the estimate for unknown active cases the 2nd of June has increased to 6964/14060. However, in one month there was a considerable reduction of unknown active cases, that now ranges from 691 to 1558. The total cases had increased ranging from 673128 to 957317. For the other regions there could be a significant bias due to the reopening of movements between regions.

**Fig. 8.**
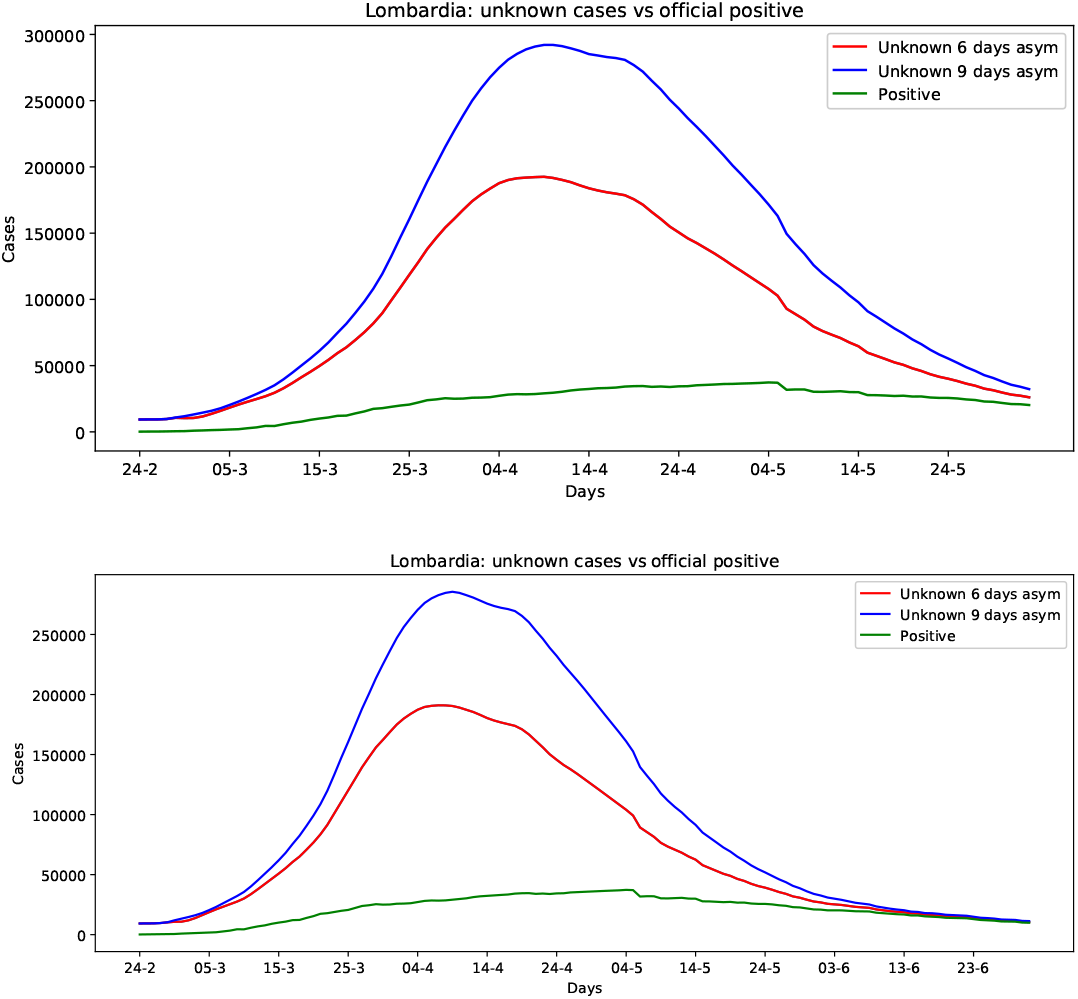
Lombardia region the 2nd of June and at the beginning of July 2020.

## 4 Conclusions

I presented the SEIAMPR system a novel simulation based model for COVID-19. The simulator provides an effective method to estimate the number of unknown cases and asymptomatic in a given area. It is based on an adaptive simulated process that is integrated with real data. SEIAMPR can be used as a tool to show the success of lockdown strategies and to design adequate exit strategies. Moreover, it allows to evaluate the role of asymptomatic in the epidemic in many settings and under different hypothesis.

The simulator can be easily used for modelling the epidemic in a given area, it only needs current positive, new positive, infected, recovered and NS time series to work, and the tuning process is simple with respect to that of mathematical models. The produced estimates can be improved, if better data are available. For example, the number of asymptomatic among the official cases, or a more accurate description of NS distinguishing the diagnostic ones with those used for recovered cases and others, like those used for screening policies on health professionals.

The limitation of the SEIAMPR model is that it cannot be used as a forecasting tool in the current setting, given that it works with real data. Adding this functionality will be the subject of future work.

A study involving 21 regions in Italy shows that the real COVID-19 cases are actually many more with respect to official ones, as confirmed by serological tests. It also highlight the effectiveness of the lockdown strategy, which obtained a considerable reduction of unknown active cases saving a lot of lives. Finally, the criticisms of region Lombardia are highlighted, with many unknown cases ranging from 6813 to 13390 still active before the 3rd of June 2020, the reopening day.

## Data Availability

The used data are available in the Italian Civil Protection site.
The source code is available upon request.

https://github.com/pcm-dpc

## Acknowledgements

The author would like to thank the Italian Civil Protection Department, and all the staff involved for providing the data of the outbreak used in this study in the Italian regions.

## Availability of data and material

This paper is avaliable as medRxiv preprint

doi: https://doi.org/10.1101/2020.07.23.20160580.

The copyright holder for this preprint (which was not certified by peer review) is the author/funder, who has granted medRxiv a license to display the preprint in perpetuity. All rights reserved. No reuse allowed without permission.

The used data are available in this site:

https://github.com/pcm-dpc.

The result of a simulation done on the 2nd of June can be ound in this site:

http://www.cs.unibo.it/~gaspari/www/covid_2_6.html.

## Code availability

The code is available upon request.

